# Seeing the light: Improving the usability of fluorescence-guided surgery by adding an optimized secondary light source

**DOI:** 10.1101/2020.03.09.20032359

**Authors:** Jonathan T. Elliott, Dennis J. Wirth, Scott C. Davis, Jonathan D. Olson, Nathan E. Simmons, Timothy C. Ryken, Keith D. Paulsen, David W. Roberts

**Author notes:** Corresponding Author: Prof. Jonathan T. Elliott, Assistant Professor of Surgery, Senior Scientist, TEC Program, Dartmouth-Hitchcock Medical Center, 1 Medical Center Drive, Lebanon, NH 03756.

## Abstract

**Background:** Tumors that take up and metabolize 5-aminolevulinic acid (5-AlA) emit bright pink fluorescence when illuminated with blue light, aiding surgeons in identifying the margin of resection. The adoption of this method is hindered by the blue light illumination, which is too dim to safely operate under, and therefore, necessitates switching back and forth from white-light mode. This paper examines the addition of an optimized secondary illuminant adapter (SIA) to improve usability of blue-light mode without degrading tumor contrast.

**Methods:** We used color science methods to evaluate the color of the secondary illuminant and its impact on color rendering index (CRI) as well as the tumor-to-background color contrast (TBCC). A secondary illuminant adapter was built to provide 475-600 nm light the intensity of which can be controlled by the surgeon and was evaluated in two patients.

**Results:** Secondary illuminant color had opposing effects on color rendering index (CRI) and tumor to background color contrast (TBCC); providing surgeon control of intensity allows this trade-off to be balanced in real-time. Experience in two cases suggests additional visibility adds value.

**Conclusion:** The addition of a secondary illuminant may mitigate surgeon complaints that the operative field is too dark under the blue light illumination required for 5-ALA fluorescence guidance by providing improved CRI without completely sacrificing TBCC.

## Introduction

Fluorescence guided surgery (FGS) is gaining popularity as a tool for improving extent of tumor resection in high grade glioma (HGG) ^1–5^. Currently, while there are a handful of approved fluorophores that can be visualized intraoperatively, 5-aminolevulinic acid (5-ALA), which is converted to the fluorophore protoporphyrin IX (PpIX) and preferentially accumulates in tumor cells, remains one of the most popular. The quick adoption of 5-ALA is attributable to three factors: 1) It can be visualized directly through compatible operating microscopes; 2) its near-monochromatic emission at 635 nm gives highly saturated reddish-purple or purplish-pink signal that contrasts strongly with tissue reflectance under blue excitation; 3) it is highly specific for high-grade glioma^5^, meningioma^6^ and metastasis^2^. Recent studies suggest 5-ALA contributes to improvements in disease free progression in HGG,^7^ and other tumors.^8–12^ Increased sensitivity of quantitative imaging methods^13–15^ may further expand the role of 5-ALA.

Interfering with the ease of use of 5-ALA by neurosurgeons, however, is the suboptimal illumination of the surgical field under blue light conditions. Difficulty visualizing fine structure under those conditions prevents many surgeons from performing the procedure while using fluorescence-guidance, relegating the technology’s role to intermittent assessment of the field, particularly at the presumed end of resection. The importance of predictable and controlled illumination during surgery has been recognized from the earliest days of modern scientific surgery—even before electricity was available, surgery was performed on the top east-facing room in the early morning to maximize the availability of natural light. Color is an interpretation, by human visual perception, of reflected or emitted light spectra governed by the object’s optical properties (scatter, absorption, and luminescence) and the illuminant.^16^ The appearance of an object is determined by both the *white-point* of the illuminant and the reflectance spectrum of the object, resulting in perceived color and lustre. Because a blue light illuminant occupies a non-overlaping locus in the visible color space, it cannot adequately render tissue color, which is heavily represented in the red, pink and yellowish-pink part of the visible color space. As a result, the latter appears dark, cold, and tissue lacks its natural visual texture due to the absence of discernable color variations; PpIX emission, however, occupies a locus along the line of purples, and appears as a vibrant (saturated) reddish-purple or pinkish-purple color, as the 635 nm emission blends with the 405 nm excitation.

Operating under blue illumination impacts surgeons’ visualization of critical nerves and other landmarks. Notably, even frank bleeding in the surgical field (an important clinical observation) is not perceivable until enough blood covers a large enough area of the field to darken the blue-light illuminated background. Despite these limitations, however, because of the intensity of color contrast between PpIX-enhancing and non-enhancing tissue, the challenges in usability are often outweighed by the marked improvements in tumor discrimination. Therefore, surgeons often switch between blue and white-light illumination manually during the course of surgery.

To overcome visual limitations of operating in blue light mode, while preserving the benefits of PpIX emission afforded to tumor identification, we introduced a secondary illuminant into the surgical field whose spectrum was optimized to achieve two competing goals: maximizing the contrast between 5-ALA-enhancing tumor cells and the background, and maximizing the amount of contamination of the excitation source and overlap with the PpIX emission peak. Specifically, we explored how the color of the light spectrum added to the blue illumination enhanced the color of non-fluorescent tissue in the surgical field (color rendering index) and affected the contrast between fluorescent tumor and non-fluorescent background. Conceptually, the approach is illustrated in **Figure 1**.

**Figure 1.**
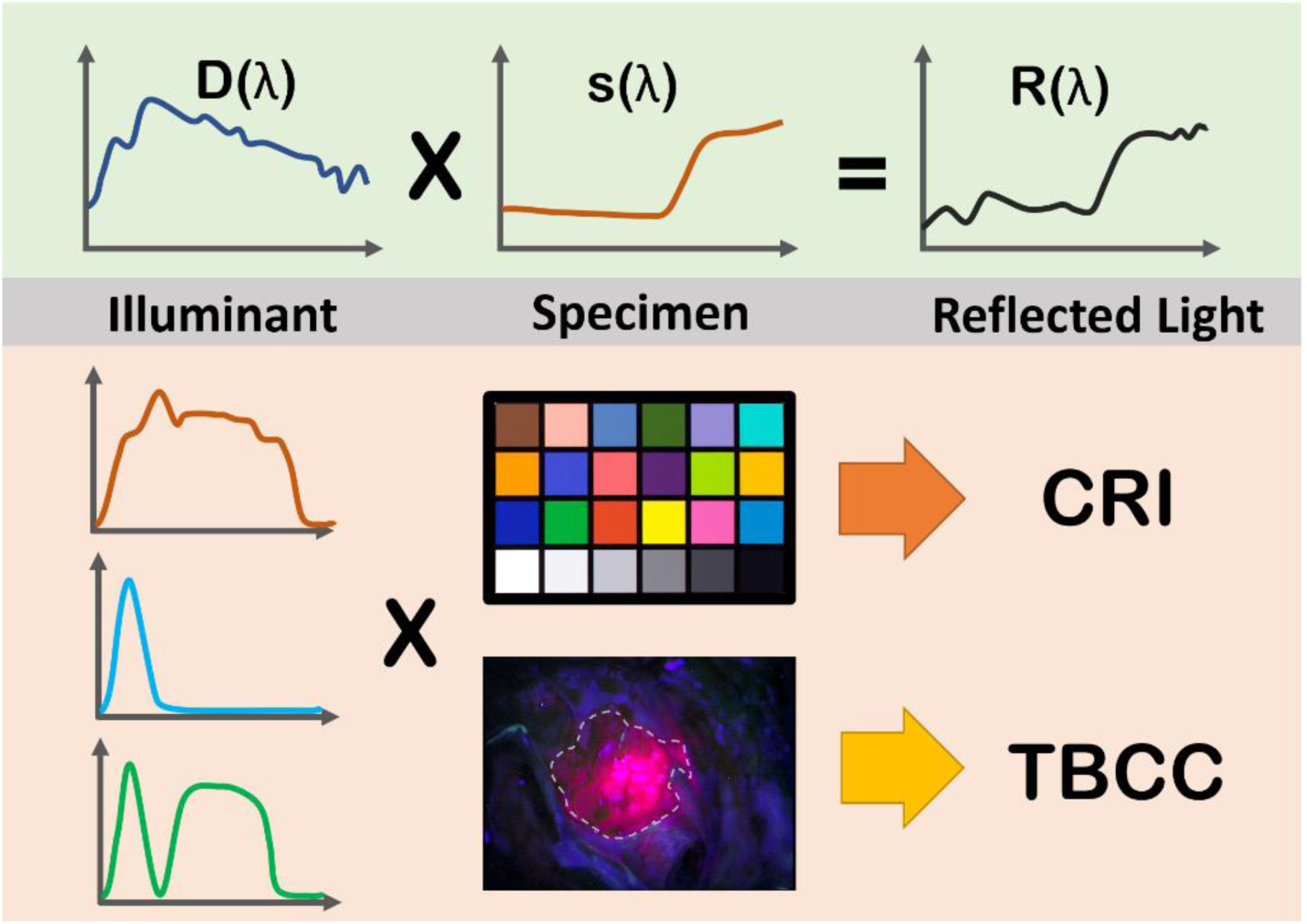
Perceived color of reflected light, R(λ), is a result of the intrinsic spectral reflectance of an object, s(λ), and the spectral density of the illuminant, D(λ). Different illuminants can result in dramatically different color renderings—measured by the color rendering index (CRI). In the context of surgical guidance, the tumor-to-background color contrast (TBCC) is also affected by the illuminant.

## Materials and Methods

### Color Theory

What we perceive as color is a result of both the intrinsic properties of an object, and the properties of the light used to illuminate it. Without light, we cannot see color, and under different types of illumination, color appears differently. The dependency of illumination on the evaluation or communication of color has led to the development of standardized illuminants such as D65, the spectral power distribution of daylight in the Northern European sky. Formally, we can define the spectrum of light reflected from the surface of a specimen, *R(λ)*, as the product of the illuminant spectrum, *D(λ)*, and the intrinsic spectral reflectance of the object, *s(λ)*. The top row of **Figure 1** summarizes this formalism. To understand the effect of the illuminant on the light reflected off the surface of a specimen, we used a standardized illuminant and hyperspectral imaging camera to obtain the pixel-by-pixel spectral reflectance of a ColorChecker card and of the surgical field from nine patients (see subsection “Clinical Study” below). These intrinsic spectra were convolved with different illuminants to obtain reflection spectra specific to these lighting conditions. To quantitatively assess these effects, two metrics were used: (1) the color rendering index (CRI), which measures the ability of a light source to render the ColorChecker test colors faithfully in comparison to the standard D65 light source,^17^ and the tumor to background color contrast (TBCC), which measures the average distance (Euclidean) between the colors in the tumor and background regions of the image. These two metrics were used in simulations to optimize the secondary light source. **Figure 1** summarizes this approach, and additional technical details are provided in the Online Content.

### Secondary Light Source Instrumentation

A Pentero BLUE400 operating microscope (Zeiss) was fitted with a custom designed and 3D printed secondary illuminant adapter (SIA), that attaches to the bottom of the microscope head via the Zeiss standard dovetail interface (Fig. ESM1, Online Content). The SIA contained a 7-W Cree XM-L T4 650LM LED emitter (SKU 1685402, FastTech.com), with color temperature range of 3700-5000 K, connected to a variable-intensity LED driver (LEDD1B, Thorlabs, Inc., Newton, NJ). Two filters (short-pass filter, SPF, and long-pass filter, LPF) were placed between the LED board to achieve the desired bandwidth of light (**Figure 2**) and a plano convex lens to expand the profile of the light. At a 300-mm working distance, when the dial on the LED driver was set to 9, maximum power meter density was measured as 14.6 mW/cm2. For the 8 cm radius spot size at 300 mm distance, the illuminance was approximately 40,000 lux.

**Figure 2.**
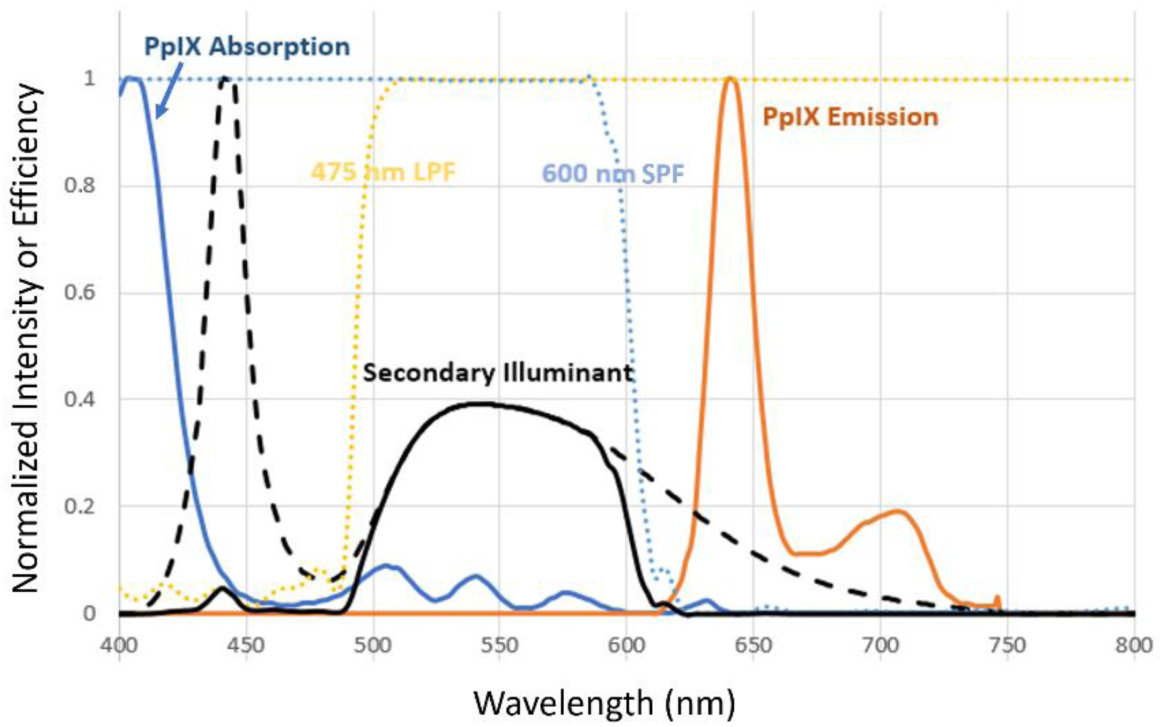
The absorption (solid blue line) and emission (solid orange line) spectra of PpIX along with the LED spectrum (dashed line) and the portion that passes through the long-pass filter (LPF, yellow dotted line) and short-pass filter (SPF; blue dotted line) forming the secondary illuminant (black line).

### Clinical Study

All study procedures were approved by Dartmouth’s IRB (Study No. 00028077; ClinitalTrials.gov NCT02191488). Nine surgical patients with operable brain tumors, presenting between September 2014 and August 2015, provided informed consent before the surgery. Three hours before surgery they were administered 20 mg/kg of ALA (DUSA Pharmaceuticals, Tarrytown, NY), used off-label under NDA 208630 since the oral route of administration was not approved in the US at the time of the study. Craniotomy was performed using navigation (StealthStationS7, Medtronic Navigation, Inc., Louisville, CO). Upon visualization of tumor and PpIX emission, sequential images under white-light and blue-light were acquired from four locations. Video and images acquired using the SIA were obtained under the same protocol following an IRB-approved planned deviation and a subsequent approved modification to include the SIA as a new non-significant risk (NSR) device. Following sequential white-light and blue-light imaging, video was acquired while the LED driver dial was turned up and down, to illustrate the changing contrast provided by the SIA.

## Results

### Characterization of Conventional Operating Microscope Illumination Modes

Thirty-six pairs of images from nine patients were obtained and characterized to determine the color gamut (*i*.*e*., the range of colors represented across images of a particular group). **Figure 3** presents the color gamuts under white light and blue light illumination within the cranial windows as probability density functions that a particular pixel will have a particular chromaticity. White light views of the surgical field are strongly weighted in the red to yellowish-pink field whereas the Blue images show a locus in the opposing side of the color space. From these chromaticity diagrams, it is evident the change in illuminant from white to blue has a dramatic effect on the reflected color, represented as chromaticity in these maps. Pixels that represent a wide range of red, pink and pinkish-yellow under a standard illuminant are compressed into the bottom left art of the chromaticity map, rendering them much less distinguishable.

**Figure 3.**
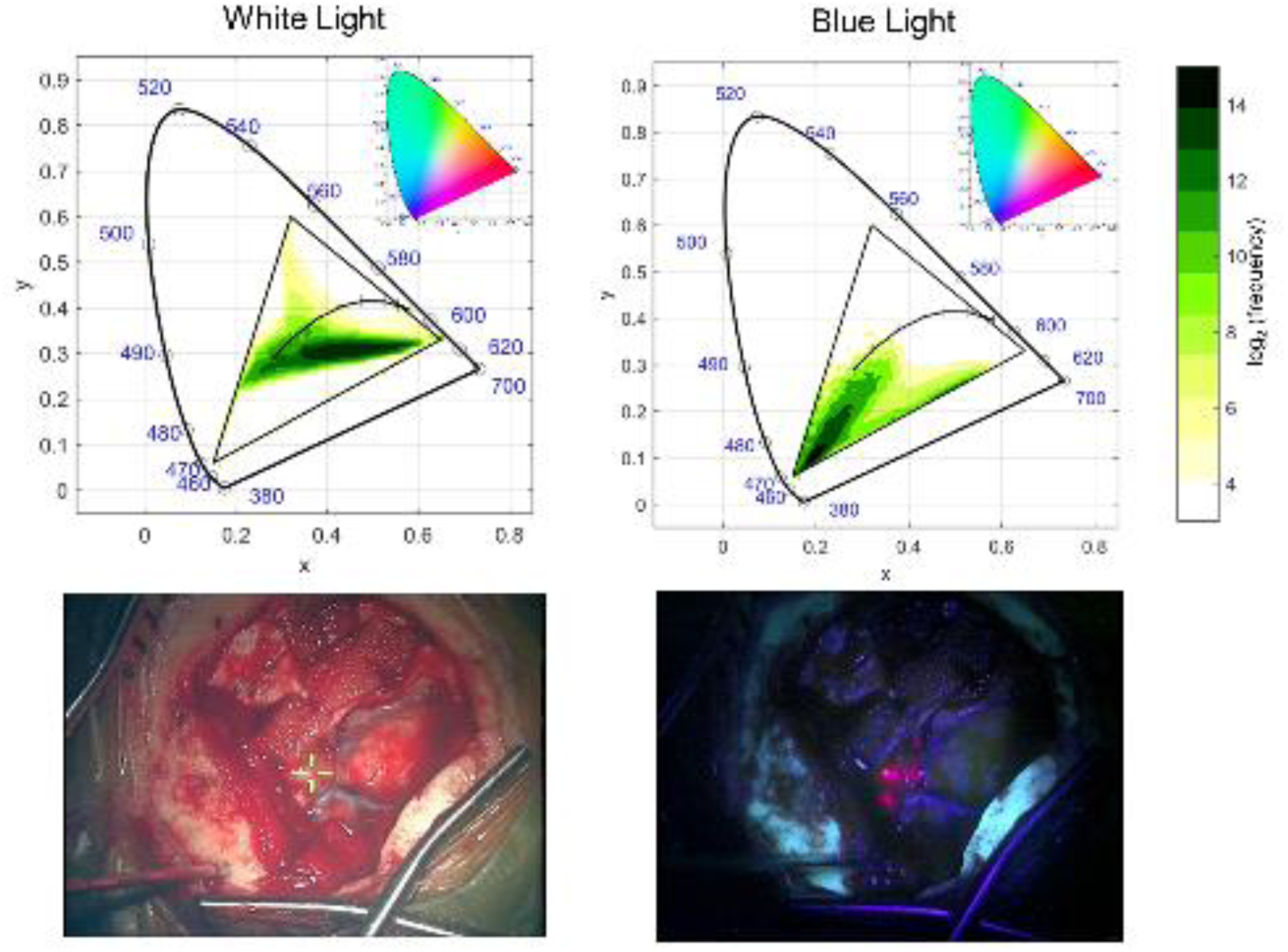
The probability density map of chromaticity calculated for thirty six pairs of intraoperative images under white-light and blue-light illumination. The top row shows CIE 1931 chromaticity diagrams with the color gamut indicated by the probability density map. Inset, a reference chromaticity map showing all visible colors is provided for reference. The bottom two images show representative white-light and blue-light images taken from the set of thirty-six pairs.

Given the dramatic impact that the choice of illuminant has on the appearance of tissue, it is therefore not surprising that the FDA has guidelines for what type of lighting is used in the operating room. **Table 1** summarizes the key characteristics of light sources used in surgery; the CIE D65 standard illuminant is included for comparison. For each characteristic, values recommended by the FDA, found in the draft IEC standard 60601-2-41/Ed1, are listed as well. The Pentero white-light is optimized to FDA guidelines with a spectrum approximating daylight (D65 CIE Standard). A hot mirror reduces transmitted light beyond 700 nm avoiding unnecessary energy deposition. The Pentero BLUE400 mode applies a 435 nm on the white-light source, while simultaneously increasing the source power by 3.5 fold to ensure adequate irradiance. The resulting monochromatic spectrum is unable to accurately render colors, and its chromaticity coordinates approach the spectral locus of the 1931 x,y chromaticity space. One of the most intuitive ways to describe a light source is by its warmth or coldness—a reference to the physics concept that black, non-reflective objects (i.e., ‘black bodies’) emit light when heated based on their temperature. As an object is heated, it’s chromaticity can be traced on the chromaticity map as an arc moving from warm to cool colors. This Planckian Locus, as it is called, intersects only red, orange yellow, white and bluish white regions of chromaticity space (**Figure 3**). Therefore, while the other three light sources have well-defined color temperatures, the saturated blue Pentero source is said to have a temperature of infinity. Given the human eye is relatively insensitive to blue light, its luminance (which is a function of both the fluence and the wavelength-dependent efficiency of the vision system) falls below the FDA guidelines for surgical lighting. Additionally, the BLUE400 mode has poor color rendering and low luminance, representing key challenges for surgeons when operating in this mode.

**Table 1.**
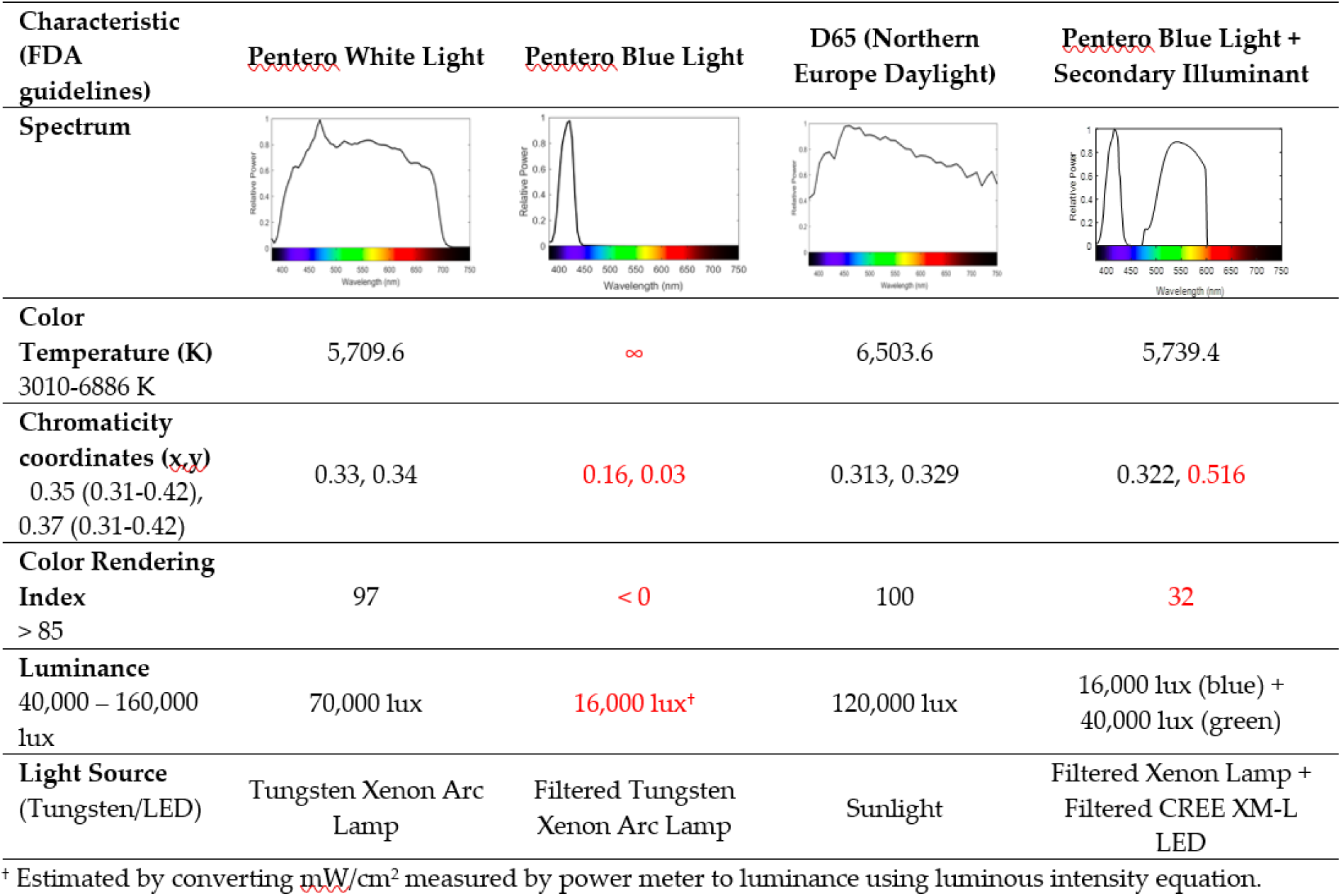
Spectral characteristics of light sources used in surgery, the D65 daylight CIE reference spectrum, and the combined light source evaluated here. Spectrum of both light sources, as measured by spectrometer. FDA guidelines for each characteristic are provided in the first column. Values outside of the guidelines are highlighted in red.

| Characteristic<br>(FDA<br>guidelines) | Pentero White Light | Pentero Blue Light | D65 (Northern<br>Europe Daylight) | Pentero Blue Light +<br>Secondary Illuminant |
| --- | --- | --- | --- | --- |
| **Spectrum** |  |  |  |  |
| **Color Temperature (K)**<br>3010-6886 K | 5,709.6 | ∞ | 6,503.6 | 5,739.4 |
| **Chromaticity coordinates (x,y)**<br>0.35 (0.31-0.42),<br>0.37 (0.31-0.42) | 0.33, 0.34 | 0.16, 0.03 | 0.313, 0.329 | 0.322, 0.516 |
| **Color Rendering Index**<br>> 85 | 97 | < 0 | 100 | 32 |
| **Luminance**<br>40,000 – 160,000 lux | 70,000 lux | 16,000 lux<sup>†</sup> | 120,000 lux | 16,000 lux (blue) +<br>40,000 lux (green) |
| **Light Source**<br>(Tungsten/LED) | Tungsten Xenon Arc Lamp | Filtered Tungsten Xenon Arc Lamp | Sunlight | Filtered Xenon Lamp +<br>Filtered CREE XM-L LED |

<sup>†</sup> Estimated by converting mW/cm<sup>2</sup> measured by power meter to luminance using luminous intensity equation.

### Effect of Secondary Illuminant Addition on Color Rendering Index and Tumor Contrast

To understand how secondary illuminant selection influences the color rendering index (CRI), a key characteristic in FDA guidelines and quantitative assessment of how well colors are rendered by an illuminant, experiments were obtained using a ColorChecker—an industry-standard card containing 24 color squares representative of the typical colors observed in the natural world. The full details of this experiment are provided in the Online Content, but the salient points are summarized here.

Hyperspectral images of the ColorChecker card (X-Rite, Inc., Grand Rapids, MI) acquired under white-light and blue-light illumination were used to evaluate the effect of varying intensity, short-pass and long-pass filter selection of the secondary illuminant. As expected, CRI decreased monotonically when the short-pass filter center wavelength was decreased, making the light more blue and narrower. The worst filter combinations occur with narrow bands of light centered around 450 nm and 575 nm. For CRI, the best bandwidth of light to add to the blue illumination spans the 475-650 nm range. In this case, the CRI approaches the FDA guideline of 85.

The tumor-to-background color contrast (TBCC) observed for different filter characteristics exhibits a more complex relationship. To understand the impact of the illuminant on tumor-to-background color, illuminants of different parameters were simulated using hyperspectral images acquired from patients undergoing glioma. **Figure 1** briefly summarizes how these simulations were performed, and once again, more details are provided in the Online Content. Patient images used to simulate the addition of hypothetical light sources are shown in **Figure 4**. Little to no impact was observed on TBCC when additional light of very cold temperature was added. However, as expected, the negative impact on TBCC was strongest when the secondary illuminant spanned the red region of the spectrum and the PpIX principle emission, causing light reflecting off the tissue to overlap with the tumor PpIX emission.

**Figure 4.**
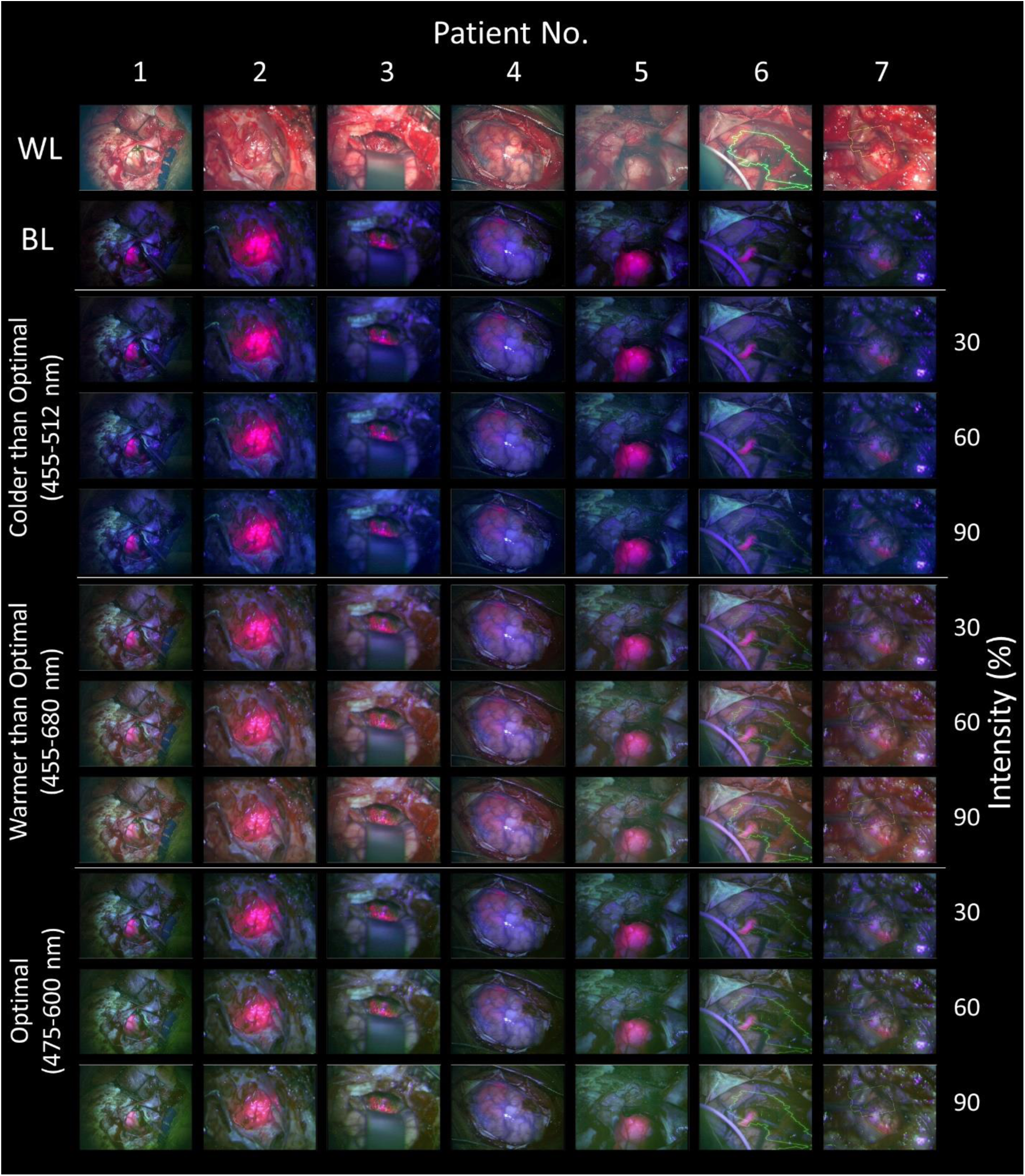
Images acquired under white-light (WL) and blue-light (BL) for seven patients undergoing craniotomy for tumor resection with the addition of secondary illuminants (simulated) having different spectral properties (cool, warm and optimal) at different intensities (3, 6, and 9).

Since these two parameters work in opposition, the overall selection of filter and intensity is not straight-forward. Since no direct means of mapping outcome to either TBCC or CRI, which would provide a weighting factor, the optimal profile remains unknown. Based on our previously described CRI and TBCC targets (CRI = 30, ΔTBCC < 15%), an optimal 475-600 nm spectral band was identified (see Fig 1). Because the trade-off will sacrifice TBCC for CRI, the best strategy may be to allow the surgeon to retain control of the intensity of the illuminant and select a secondary source with better CRI, which will provide the flexibility needed for enhancing tumors which show a wide variability in the 5-ALA signal intensity.

### Video Acquired During Fluorescence Guided Surgery using an Optimized Secondary Illuminant

Two patients enrolled to receive 5-ALA FGR were imaged under blue light and white-light illumination during the craniotomy procedure. These images were recorded under the same conditions (white-light power of 30%, blue-light power of 100%, distance of 300 mm and magnification of 1.6x) as acquisitions of the ColorChecker card (see ColorChecker Card Accuracy in Materials and Methods). The optimized secondary illuminant, constructed with a 475 nm LPF and 600 nm SPF, was then used to illuminate the surgical field during BLUE400 mode. Video showing the transition between BLUE400 and BLUE400 + secondary illuminant (SI) was recorded (Online Content), and a panel of screen captures from the video are presented in **Figure 5**.

**Figure 5.**
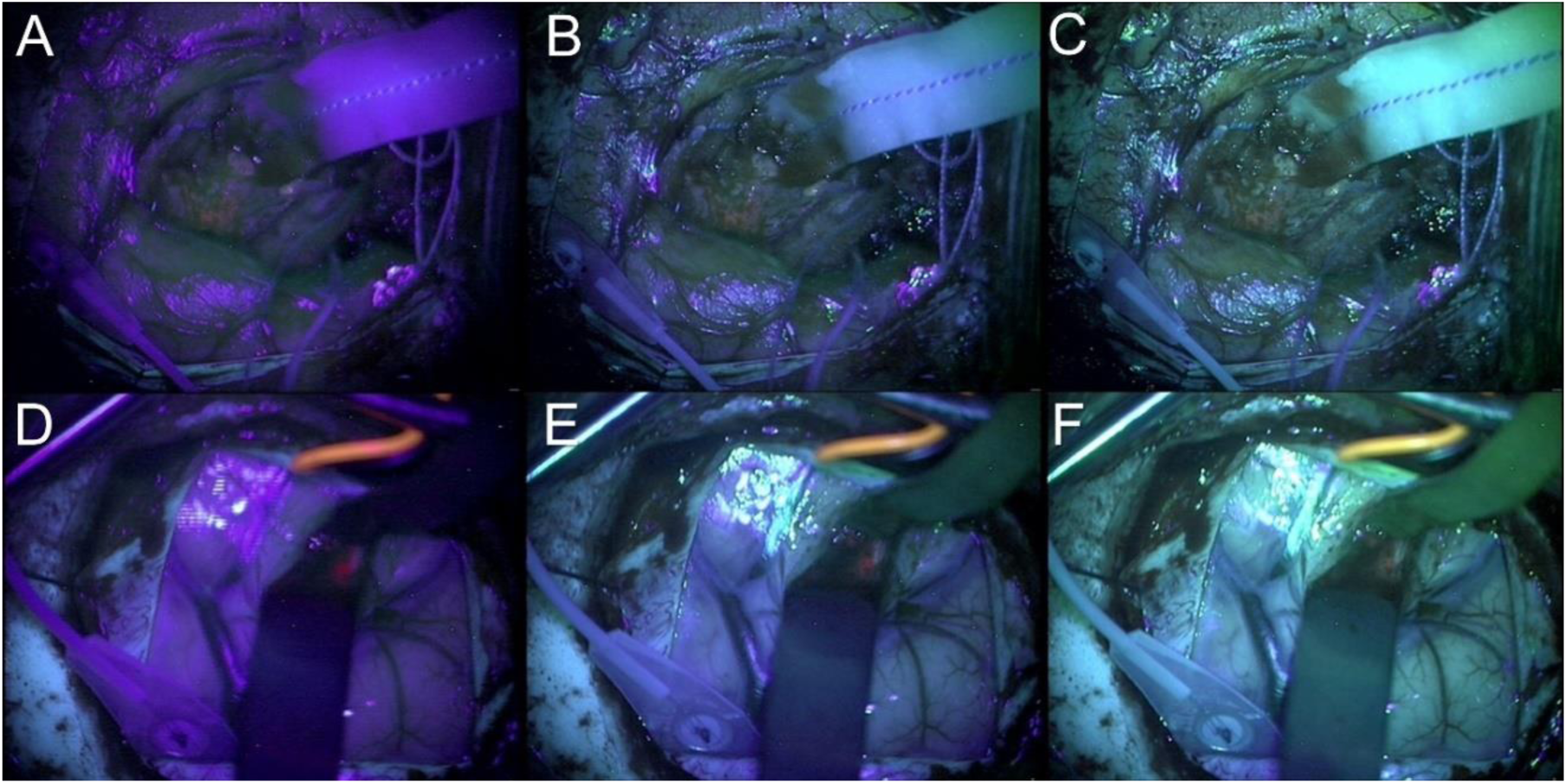
Link to video. Images acquired during 5-ALA FGS from patient one (top row) and patient two (bottom row) under blue light only, blue light with low secondary illumination, and blue light with medium secondary illumination (left to right).

We set the maximum power of the secondary illuminant to 3x the irradiance of the blue light source (48,000 lux). During deployment of the technology in the OR, about 30% of the maximum LED power was used in the two patient cases summarized in **Figure 5**. In these surgeries, tumor exhibited lower levels of PpIX fluorescence than in many of the cases shown in **Figure 4**; and therefore, they may not represent conditions under which a secondary illuminant would provide the most added value. Nevertheless, the surgeon (T.R.) reported that the secondary illuminant was able to highlight features that were otherwise obscured in BLUE400 mode. This is observed in the video showing the transition from white light to blue light to blue light with the SIA (**Video 1**, Online Content). In highly fluorescent tumors, most of the surgery could be performed with the secondary light source on, although PpIX fluorescence would only be visible if a carefully shaped spectrum was used.

## Discussion

The salient finding of this paper is that, when considering the selection of a secondary illuminant to improve the color rendering of the surgical field, the negative effect on tumor-to-background color contrast is unavoidable. There is currently no outcomes data that would provide the impact of either CRI improvement or TBCC degradation on disease-free progression, so no universal optimization is possible. A secondary objective of this paper is to provide a framework to consider the impact of illumination on appearance of tissue during surgery.

Given no global optimum exists across both CRI and TBCC, cost term weighting will impact the overall optimization. In the absence of clinical outcome data—*e*.*g*., how CRI influences avoidance of critical structures like nerves and vessels, or how TBCC degradation impacts extent of tumor resection— the weighting is arbitrary. Notwithstanding, we selected a CRI region corresponding to 30 for the purpose of this study, which represents a light source with a color rendering quality better than a high-pressure sodium light but worse than a fluorescent light. While these CRI values are below FDA guidelines, they are common illuminants in day-to-day life. Determining the decrease in TBCC that would be tolerated is more arbitrary. The largest decreases in contrast occurred when overlapping with the emission peak of PpIX. Outside of this region, contrast was greatly improved. The selected filter-set produced about 15% decrease in contrast. Once the filters were selected, the intensity of the light was varied, allowing the surgeon to enhance contrast or the color rendering of the operative field as needed. It should be emphasized that the “on the fly” adjustment can eliminate any degradation in TBCC by turning off the secondary illuminant, but can only enhance CRI to a maximum of the CRI of the secondary light source itself. Nevertheless, the ability to fine-tune the balance between improved CRI or improved TBCC therefore allows individual surgeons to improve usability of 5-ALA, *e*.*g*., at different phases of resection within the same case, or for different cases where the concentration of 5-ALA induced PpIX fluorescence might be bright enough to permit more secondary illumination. In this sense, the primary objective of this paper to improve the usability of 5-ALA surgical guidance was achieved.

In a recent publication, Schwake et al. presented data where 5-ALA and fluorescein were used simultaneously, remarking that the principle benefit of fluorescein was background illumination facilitating visualization of the eloquent brain.^18^ The SIA would theoretically provide some of the same benefits without the added task, cost and risk of administering another contrast agent. While its intensity and spatial distribution could be controlled more effectively, a ‘frontlit’ approach like the SIA is more likely to produce distracting specular reflections compared with the ‘backlit’ source provided by fluorescein. It may also be argued that once the surgeon can effectively control the intensity of the SIA, why not use a white-light SIA. Evidence from studies of human perception and cognition, suggest smooth transition between BLUE400 and BLUE400+SI is more desirable than switching directly to white-light. The ability to mentally integrate information from one representation of a scene into another, a type of spatial reasoning, relies on qualitative aspects of the rendering (patterns, points of reference, landmarks) to persist even when quantitative aspects of the image (chromaticity, brightness, object saliencies) change 18. The BLUE400 + SI mode may provide a rendering of the scene with more persistent aspects relative to the BLUE400 mode, and therefore, may facilitate integrative spatial reasoning. The next step in this technology development is to compare white-light and green-light SIAs to confirm whether there is an added benefit to spatial reasoning.

## Conclusion

This proof-of-concept paper demonstrates the use of a secondary illuminant adapter, affixed to the bottom of a standard Zeiss Pentero operating microscope, to improve tissue color rendering during 5-ALA FGR. The optimized light source balanced improvements in color rendering with reduction in 5-ALA tumor-to-background contrast. Video of 5-ALA FGR acquired in two patients using the SIA is shown to demonstrate the resulting effect on color rendering and contrast. While no global optimization was possible since CRI and TBCC work in opposition when an SIA is added, the ability to allow surgeons to individually, within the context of the phase of resection or patient-specific fluorescence properties, modulate between enhanced TBCC and enhanced CRI provides substantive improvements in usability.

## Data Availability

Data is available upon request, pending a data sharing agreement approved by the IRB.

## Declarations

### Availability of data and material

upon request

### Code availability

upon request

### Ethics approval

Dartmouth IRB Study No. 00028077

### Consent to participate

Written consent was obtained from all subjects.

### Consent for publication

All authors consent to the publication of this work.

## Acknowledgements

External funding for this work was provided by NIH/NCI R00 CA190890 and NIH/NCI RO1 NS052274. The authors would also like to acknowledge internal funding support from the Dartmouth-Hitchcock Medical Center’s Department of Surgery and the Translational Engineering in Cancer Program at the Norris Cotton Cancer Center.

